# Rates of Event Capture of Ambulatory Video EEG

**DOI:** 10.1101/2022.11.13.22282197

**Authors:** Ewan S. Nurse, Timothy Hannon, Victoria Wong, Kiran M. Fernandes, Mark J. Cook

**Author notes:** Contact Prof. Mark Cook, Address: 404, Clinical Sciences Block, St Vincent’s Hospital Melbourne, 41 Victoria Parade, Fitzroy VIC 3065, Australia.

## Abstract

**Objectives:** Recording electrographic and behavioral information during epileptic and other paroxysmal events is important during video EEG monitoring. This study was undertaken to measure the event capture rate of an ambulatory service operating across Australia using a shoulder-worn EEG device and telescopic pole-mounted camera.

**Methods:** Neurologist reports were accessed retrospectively. Studies with confirmed events were identified and assessed for event capture by recording modality, whether events were reported or discovered, and wakefulness.

**Results:** 6,265 studies were identified, of which 2,788 (44.50%) had events. A total of 15,691 events were captured, of which 77.89% were reported. The EEG-ECG amplifier was active for 99.83% of events. The patient was in view of the camera for 94.90% of events. 84.89% of studies had all events on camera, and 2.65% had zero events on camera (mean=93.66%, median=100.00%). 84.42% of events from wakefulness were reported, compared to 54.27% from sleep.

**Conclusion:** Event capture was similar to previously reported rates from ambulatory studies, with higher capture rates on video. Most patients have all events captured on camera.

**Significance:** Ambulatory monitoring is capable of high rates of event capture, and the use of wide-angle cameras allows for all events to be captured in the majority of studies.

**Highlights:**

- A review was undertaken of an Australia-wide ambulatory video-EEG monitoring service
- Patients were in view of camera for 94.90% of events, and 84.89% of studies had all events on camera
- 84.42% of events from wakefulness were reported, compared to 54.27% from sleep

## 1 Introduction

Ambulatory video-EEG is increasingly being utilized in the diagnosis and management of epilepsy and other paroxysmal disorders (Primiani et al., 2020; Seneviratne and D’Souza, 2019; Tatum et al., 2022b). Ambulatory monitoring provides a resource and cost-efficient alternative to inpatient monitoring (Schomer, 2006; Slater et al., 2019; Tatum et al., 2018), particularly in the context of the ongoing COVID-19 pandemic which has significantly limited the rates of inpatient admission to epilepsy monitoring units (Ahrens et al., 2022; Beniczky et al., 2021).

The yield of ambulatory monitoring is highly variable between service providers (both in terms of percentage of studies with events and the diagnostic rate), however this is likely strongly influenced by different study populations and the duration of monitoring (Primiani et al., 2020; Syed et al., 2019). A recent study by Schulze-Bonhage et al. demonstrated that even well-established inpatient epilepsy monitoring units can have markedly different yields of events, assumedly due to duration of monitoring, patient populations (particularly the inclusion of pediatric patients) and seizure provocation methods contributing to different rates of event capture (Schulze-Bonhage et al., 2022). A significant concern in the use of ambulatory monitoring is the ability of the equipment to suitably capture the required electrographic and behavioral information outside of a clinically supervised environment (Brunnhuber et al., 2020; Goodwin et al., 2014). Hence, an improved understanding of the event capture rate of ambulatory video-EEG is warranted.

This study presents the outcomes of a retrospective service review of an ambulatory video-EEG service operating in Australia. A novel shoulder-worn EEG-ECG amplifier was used in combination with a camera mounted on a telescopic pole for ambulatory video-EEG monitoring. Data are captured and displayed on a cloud-based system, allowing for remote device troubleshooting and review. Records from 6,265 consecutive study reports yielding 15,691 events were assessed for technical outcomes, rate of event capture on camera, and rate of event capture in different behavioral states, to better understand the technical yield of ambulatory video-EEG monitoring.

## 2. Methods

### 2.1 Participants

This study was conducted under approval from the Human Research Ethics Committee of St. Vincent’s Hospital Melbourne (042/18). All participants provided written, informed consent. Clinical records of patients referred for ambulatory video EEG were accessed retrospectively. All records were stored in a secure cloud platform allowing for automated analysis.

Patients were referred for ambulatory video EEG monitoring, typically for differential diagnosis, assessment of treatment efficacy, or characterizing seizure types. All monitoring was undertaken in Australia. Patients would attend a clinic for the fitting of the EEG-ECG device, and to receive instructions for how to operate the camera system. Data from 24 clinics was included in this analysis. Patients would then return to their residence for the duration of monitoring. Monitoring duration ranged from 1 to 7 days. No sleep deprivation or drug tapering were undertaken during monitoring. All equipment used is manufactured by Seer Medical Pty Ltd (Melbourne, Australia), and is CE, FDA, and TGA listed.

### 2.2 Video-EEG-ECG system

The EEG-ECG amplifier is worn on the shoulders, and weighs 225g. A single lead connects the amplifier to the analog/digital converter at the top of the head (centered at approximately CPz). The analog/digital converter connects to the electrodes via an array of pogo-pins. 21 channels of EEG are recorded from 1cm diameter Ag/AgCl disposable electrodes placed in the 10-20 system, which are affixed with a polymer adhesive and conductive gel (Nurse et al., 2022). Three ECG electrodes are placed on the chest. EEG and ECG are sampled at 500 Hz, 24 bits per sample. The batteries are stored in the inferior ends of the system, sitting approximately on the collar bone. The battery life is approximately 10 days, and hence do not need to be changed during a standard recording. Input impedance is >500MOhms, and Common Mode Rejection Ratio is >100dB. This connects wirelessly to the camera via Bluetooth 5 or WiFi. These technical specifications meet the recently proposed technical standards for ambulatory EEG (Tatum et al., 2022a)

The camera sits upon a telescopic pole, with the center of the camera 1800mm from the ground at full height. Camera resolution is 2560×1920 pixels, sampling at 30 frames per second. The lens captures approximately 180-degree field-of-view and has a built-in microphone. In low light conditions, the camera transitions to infra-red night vision. Without mains power, the video can record for approximately 4 hours. The camera and amplifier stream securely to a cloud platform via 4G, WiFi, or ethernet connection. Patients are instructed to move the camera system with them as they move about their environment, with the exception of going to the bathroom, or other short periods away from the camera.

Patients are provided with a tablet device that runs an application allowing them to connect the system to their home WiFi, informs if the system is recording data, and can display impedance checks (triggered remotely by a technician) to facilitate electrode repair. Technicians periodically monitor the system remotely to ensure data is being recorded, and signal quality is maintained.

### 2.3 Event reporting

Patients or caregivers record events via a mobile seizure diary app (Seer Health, 2022). Information recorded includes the time the log was made, the time the event occurred, the estimated duration of the event, and a free-text field for describing symptoms. The timing of the events is synchronized to the video-EEG data.

After the recording is completed, the data from each study was reviewed by a neurophysiologist. Each event from the recording is classified as:

- If the amplifier was active
- If the audio recorder was active
- If the video recorder was active
- If the patient was in view of the video
- If the event was reported, or discovered by clinical staff
- If the patient was awake, drowsy, or asleep at the onset of the event Events may be either reported by the patient/carer or discovered on review of the recording (a discovered event). The recording is viewed in a browser-based video-EEG-ECG reviewing/reporting software package.

### 2.4 Analysis

All studies since the implementation of this clinical reporting system in March 2020 to July 2022 were included in this analysis. The percentage of events with different recording modalities and clinical characteristics was assessed. Furthermore, the fraction of events in view of video on a per-study basis were analyzed, as this has been previously identified as a drawback of ambulatory monitoring (Goodwin et al., 2014). All data retrieval and analysis was undertaken in Python v3.8.2.

## 3. Results

### 3.1 Event yield

A total of 6,265 studies were identified. Of these, 2,788 (44.50%) had events (either reported or discovered). A total of 15,691 events were captured, comprising of 12,221 reported events and 3,470 discovered events (77.89% reported, 22.11% discovered).

The amplifier was active for 99.83% of events. Note this makes no qualitative assessment of the EEG or ECG trace, only that the amplifier was powered on and recording. The video recording was active for 98.15% of events, and audio for 97.15%. The patient was in view of the video for 94.90% of the total number of events. The patient was in view for 94.06% of all reported events, and 96.37% of discovered events. The patient was in view of the video and the amplifier was active in 94.45% of all events, and 94.43% of all events also had audio available. These values are presented in Table 1.

**Table 1:**
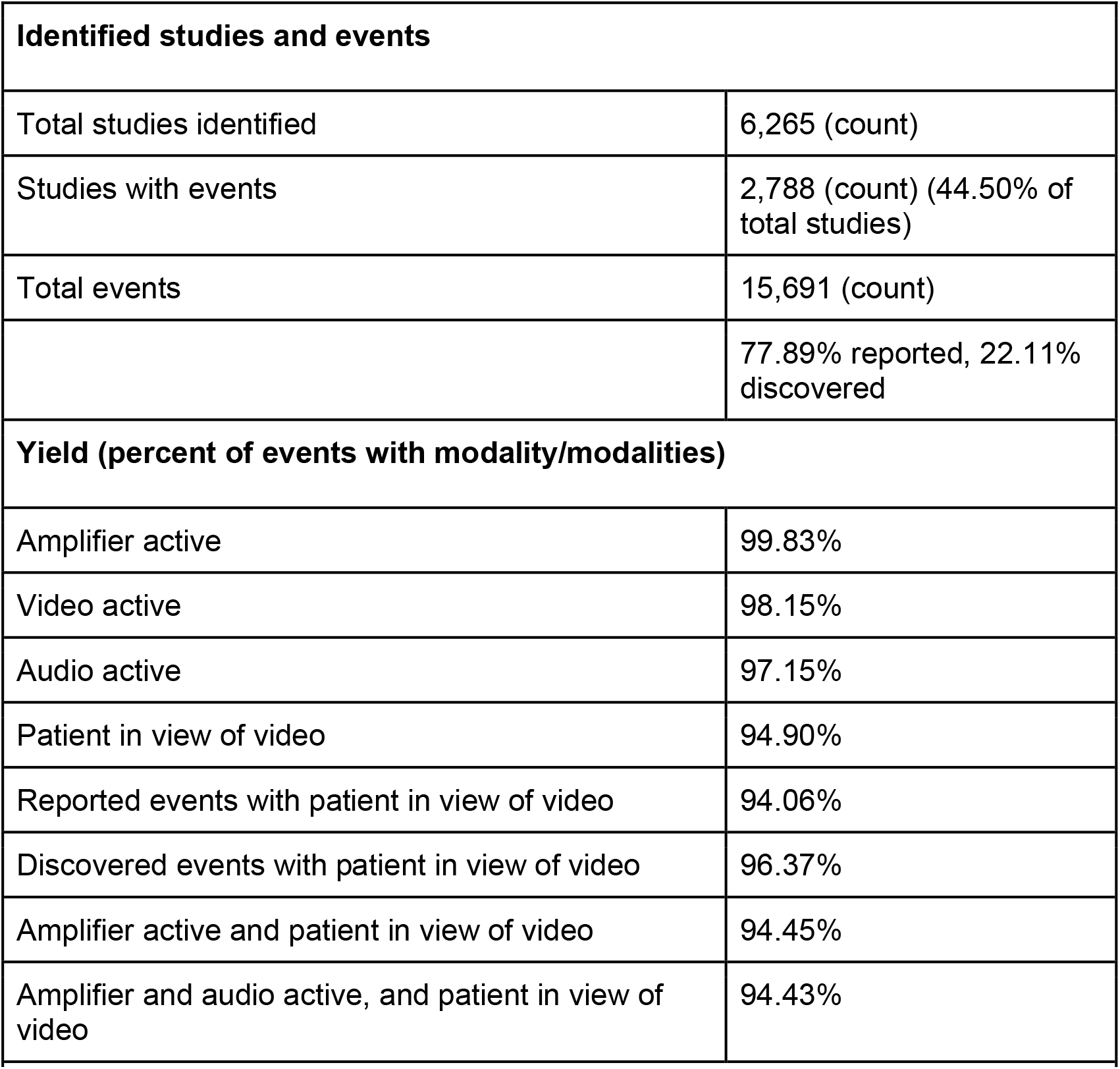
Event yield and rates of event capture for different data modalities during ambulatory vEEG.

### 3.2 Events on camera per study

For each study with identified events, the proportion of events that occurred on camera was calculated. This is displayed as a histogram in Figure 1. 84.89% of studies had all events captured on camera. 96.08% had at least half of all events on camera. 2.65% of studies with events had no events captured on camera. Hence, the vast majority of studies capture the vast majority of events on camera.

**Figure 1:**
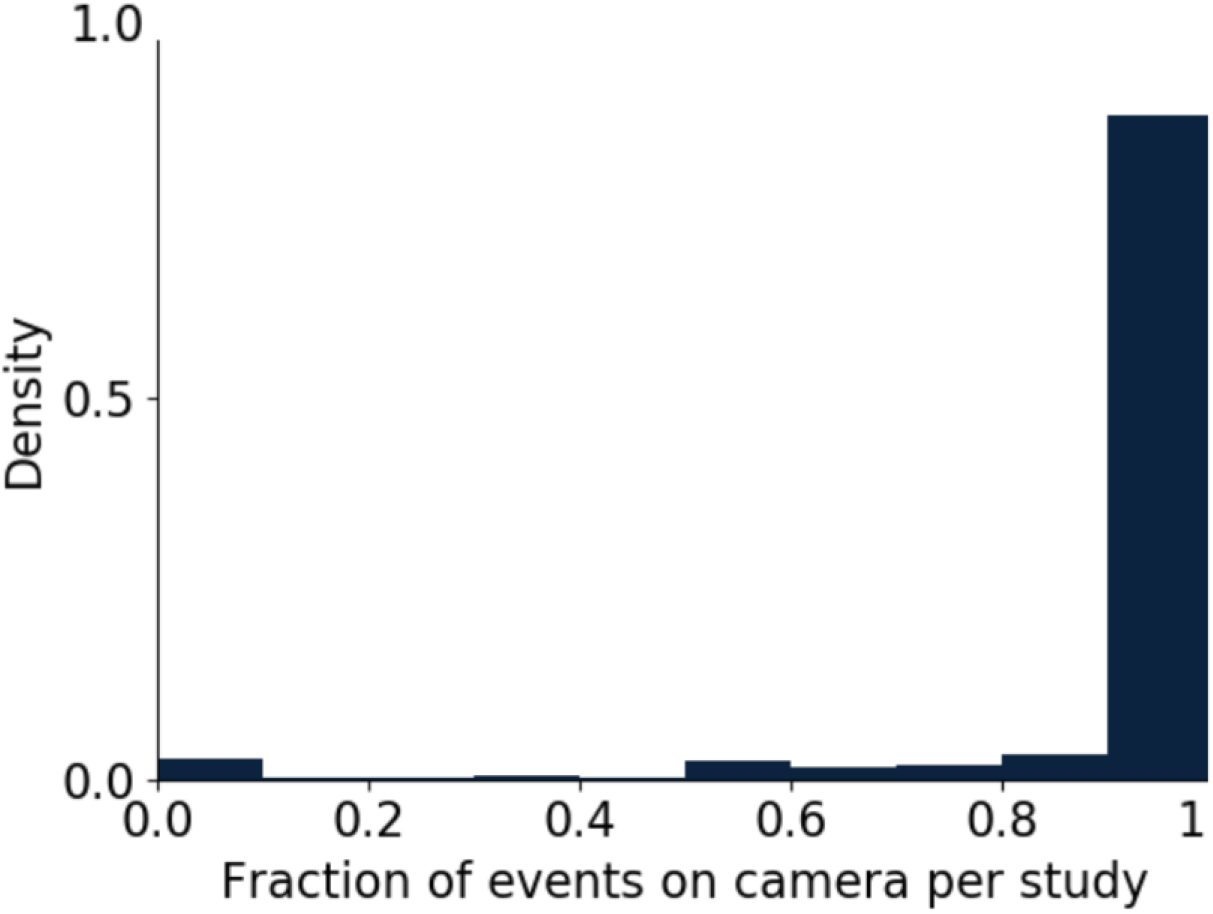
Density histogram of fraction of event capture on video per study. 1 on the x-axis represents all events in a study were captured on camera, and 0 represents none of the events were on camera. 96.08% of studies have at least half of events captured on camera. 84.89% of studies have all events captured on camera. 2.65% of studies have none of the events that occurred captured on camera. Mean = 93.66% of events captured on camera, median =100.0% of events captured on camera.

### 3.3 Patient wakefulness and event capture

79.41% of the total events were captured during wakefulness, 3.28% from drowsiness, and 17.30% from sleep. Of the events captured during wakefulness, 84.42% were reported. From drowsiness 69.47% of events were reported, and from sleep 54.27%. As might be expected, far less events from sleep are reported compared to those that begin in wakefulness. 94.12% of events from wakefulness were in view of the video. From drowsiness and sleep, the rates were 97.26% and 97.12%, respectfully. The higher rate of event capture from sleep may be expected as patients will not be moving around their environment. These results are summarized in Table 2.

**Table 2:** Patient wakefulness and rates of event capture.

| Event onset wakefulness |  |
| --- | --- |
| Awake | 79.41% |
| Drowsy | 3.28% |
| Asleep | 17.30% |

| Event onset wakefulness and reporting |  |
| --- | --- |
| Awake and event reported | 84.42% |
| Drowsy and event reported | 69.47% |
| Asleep and event reported | 54.27% |
| Event onset wakefulness and patient visibility |  |
| Awake and patient in view of video | 94.12% |
| Drowsy and patient in view of video | 97.26% |
| Asleep and patient in view of video | 97.12% |
| **Table 2: Patient wakefulness and rates of event capture** |  |

The distribution of events by hour of day is shown in Figure 2. More events are captured in waking hours (peaking at 8AM and 5PM) than the night (lowest point at 3AM).

**Figure 2:**
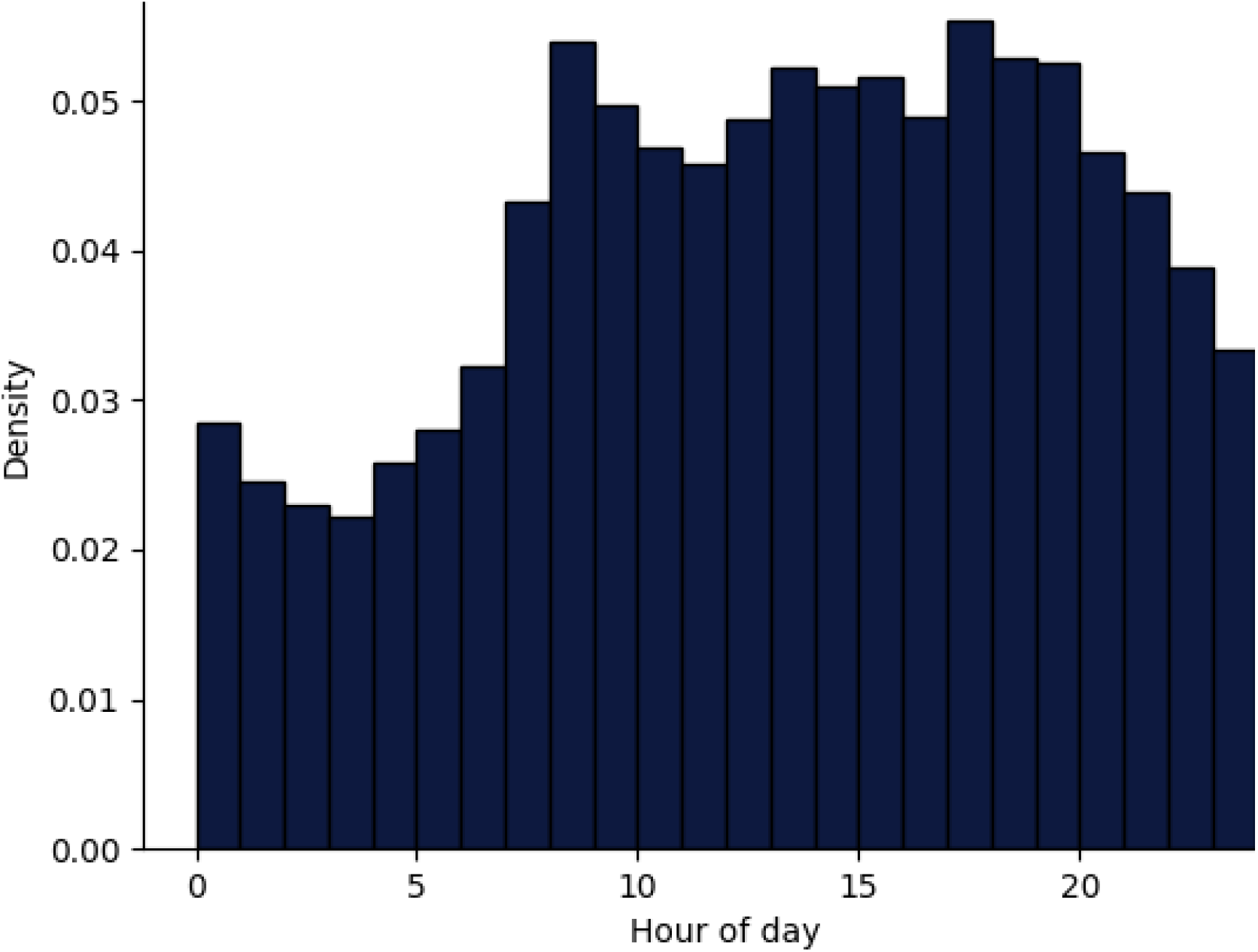
Density of events by hour of day (24h time). It can be seen that more events occur during daytime hours, and less during the night. The time with the lowest yield is 3AM, and the highest yield occurs at 8AM and 5PM.

## 4. Discussion

This study reports on the technical yield of an ambulatory video-EEG-ECG service operating across Australia from a population of patients with epileptic and/or non-epileptic events. Results from this work demonstrate that the amplifier and video were available for the vast majority of events (both reported and discovered), and that the patient is in view of the camera for the vast majority of events. As perhaps expected from this cohort, more events occurred during wakefulness, and a greater percentage of events during wakefulness were reported compared to sleep. More events occurred on camera during sleep than during wakefulness, presumably due to the patient not moving out of field-of-view while sleeping. Peaks in event rates occurred at approximately 8am and 5pm. As this cohort is composed of not only people with epilepsy but any patient referred to the service, the underlying distributions that make up these peaks are difficult to speculate on, however the sleep/wake cycle is noticeably modulating the likelihood of events.

Primani et al. previously demonstrated that the yield of events between ambulatory EEG studies can vary markedly (range 55-85%). Our work has a lower rate of event capture than the previously presented works, at 44.5%. This is however very similar to the yield of epileptic patients in a recently published comparison of two established European epilepsy centers (Schulze-Bonhage et al., 2022). The inability to withdraw ASMs or undertake other seizure provocation methods due to safety concerns will likely mean that ambulatory yield will generally remain lower than inpatient yield. Methods such as seizure cycle analysis may allow for more optimal monitoring periods to be selected when event risk is higher (Karoly et al., 2021, 2018), increasing the study yield. Direct comparison of the event rate in our study to other video EEG systems is difficult, due to a heterogeneity in the literature of inclusion criteria, definition of events and outcomes, duration of monitoring, and equipment used.

The percentage of reported events (77.89%) is much higher than previously published inpatient data including only confirmed epilepsy patients (55.5%) (Hoppe et al., 2007). This difference would largely be due to the inclusion of non-epileptic events in this work, which may be entirely subjective in nature. Events in this work were also logged via a mobile app, which may affect yield compared to a physical push-button. Similarly, the percentage of events from sleep that are reported is much higher in this work (54.27%) compared to previously reported epileptic inpatient results (14.2%) (Hoppe et al., 2007). Future work should focus on the epileptic sub-population of this cohort to identify if reported event rates are more similar to inpatient figures.

Rate of event capture on video in previous ambulatory video EEG works has been approximately 80% (Kandler et al., 2017; Primiani et al., 2020), compared to 94% in this work. This may be due to improvements in camera technology, allowing for wide-angle video capture, and automatic transitions from daylight to low-light (infrared) recording modes. The telescopic pole system may also increase the yield, as systems relying on the placement of the camera on a flat, elevated surface (i.e. a bookshelf) may not have optimal placement in all environments. Video systems with multiple cameras warrant further investigation to minimize the fraction of missed events.

This retrospective work has several limitations. Yield has not been stratified by referral type or diagnostic outcome, which would have required substantially more manual data annotation for this dataset. When equipment hasn’t captured events, we have not assessed the reason why this is the case (i.e. failure, low data quality, device not charged, etc.).

Events have only been assessed by individual clinical scientists, then reported by an individual neurologist, but assessment by multiple clinicians may improve the quality of the outcome measures. Future work should assess the rates of event capture by those others than the patient being monitored (i.e. family members) compared to the patient alone, particularly during sleep.

There currently is not a well-accepted standard for when a reported event is considered “correctly” reported when there is an electrographic or behavioral correlate. A clear definition used by the broader epilepsy community will be necessary as there is an increasing clinical use of other ambulatory monitoring devices such as chronic EEG, nocturnal monitoring devices, and wearable systems which would benefit from a standardized measure (Duun-Henriksen et al., 2020; Stirling et al., 2021; Armand Larsen et al., 2022; Brinkmann et al., 2021; Elger and Hoppe, 2018).

## 5. Conclusion

Ambulatory video EEG is increasingly being utilized in the diagnosis and management of seizure disorders. As an important diagnostic clinical tool, concerns exist around the event capture rate of ambulatory systems, as close supervision of patients becomes more difficult with remote, long-term monitoring. This work has demonstrated that the rate of event capture from a shoulder-worn EEG system combined with a camera mounted on a telescopic pole is similar to previous reports, with particularly high rates of event capture on camera. The vast majority of patients have all of their events captured on camera. Further work is required to understand the failure mechanisms of missed events, and further elucidate the mechanisms of non-reported events in the epilepsies as well as other seizure disorders.

## Data Availability

All data produced in the present study are available upon reasonable request to the authors

## Declaration

ESN and MJC declare a financial interest in Seer Medical Pty. Ltd.

We confirm that we have read the Journal’s position on issues involved in ethical publication and affirm that this report is consistent with those guidelines.

## Acknowledgements

The authors thank Drs. Dulini Mendis and Ryan Mills for their assistance with data access for this study. We also thank Tim Nelson and Thilini Perera for their discussions around the clinical and technical aspects of this work.

